# Prevalence of persistent symptoms in children during the COVID-19 pandemic: evidence from a household cohort study in England and Wales

**DOI:** 10.1101/2021.05.28.21257602

**Authors:** Faith Miller, Vincent Nguyen, Annalan MD Navaratnam, Madhumita Shrotri, Jana Kovar, Andrew C Hayward, Ellen Fragaszy, Robert W Aldridge, Pia Hardelid, on behalf of the Virus Watch Collaborative

**Author notes:** Virus Watch Collaborative: Susan Michie, Linda Wijlaars, Eleni Nastouli, Moira J Spyer, Ben Killingley, Ingemar Cox, Vasileios Lampos, Rachel A McKendry, Tao Cheng, Yunzhe Liu, Anne M Johnson, Jo Gibbs, Richard Gilson, Cyril Geismar, Sarah Beale, Isobel Braithwaite, Thomas E Byrne, Wing Lam Erica Fong, Parth Patel, Anna Aryee, Alison Rodger.

## Abstract

Using data from 4678 children participating in VirusWatch, a household cohort study, we estimated the prevalence of persistent symptoms lasting ≥4 weeks as 1.7%, and 4.6% in children with a history of SARS-CoV-2 infection. Persistent symptom prevalence was higher in girls, teenagers and children with long-term conditions.

## Introduction

An increasing number of studies are reporting that individuals who have been infected with SARS-CoV-2 may experience persistent, post-acute symptoms, which may last for many months.^1^ The majority of studies of these persistent symptoms, frequently termed ‘long COVID’, have been conducted in adults. Compared to adults, children have been much less severely affected by COVID-19 disease, and it is unclear to what extent children are affected by persistent SARS-CoV2-related symptoms.^2^ Understanding the frequency and severity of persistent symptoms following SARS-CoV-2 infection in children is crucial for informing COVID-19 paediatric vaccination policy. Two UK community-based studies have reported the prevalence of persistent symptoms (lasting >4 weeks) following SARS-CoV-2 infection in children as 4.4% using data from a symptom app,^3^ and 9.8% to 13.0% (depending on age group) in a household survey.^4^ Generally, the estimated prevalence of persistent symptoms in children with SARS-CoV-2 infection presenting in secondary care have been higher, ranging from 8% to 25% 3-7 months after infection onset.^5 6^ We estimated the prevalence of persistent symptoms reported in children during the summer, autumn and winter of 2020-2021, and their association with SARS-CoV-2 infection, in a large household-based community cohort study.

## Methods

We used data from VirusWatch, a household cohort study in England and Wales. Study design and recruitment are described elsewhere.^7^ Briefly, households were recruited starting in mid-June 2020 via a number of methods, including postcards or letters sent to the home address, social media and SMS. As of mid-March 2021, 47,813 individuals in 23,059 households had registered to take part. To participate, a household required internet access and an email address, and at least one household member had to speak sufficient English for survey completion. Participating households completed online weekly surveys (reporting a wide range of symptoms and SARS-CoV-2 swab test results), and monthly themed topic surveys. Parents consented on behalf of their children if children were <6 years old. VirusWatch also included a programme of nasopharyngeal swab sample collection, and blood collection via venepuncture or fingerprick sampling in a subset of 10,000 participants (See appendix 1). We used data from the 3^rd^ monthly survey (distributed on the 17^th^ February 2021), which asked about persistent symptoms (the ‘long covid survey’).

All children aged ≤17 years at enrolment who had either a) answered the question about persistent symptoms in the 3^rd^ monthly survey, or b) whose household had participated in at least 3 weekly surveys in a 5-week period, before the 20^th^ January 2021, were included in our analysis.

In the long covid survey, participants were asked: “*In the last year (since February 2020) have any of the household members experienced any new symptoms that have lasted for four or more weeks even if these symptoms come and go, and that are not explained by something else (eg, pre-existing chronic illness or pregnancy)?”* Participants who responded ‘*yes’* could also report the nature of the symptoms and the date of onset and resolution for the three most severe symptoms. We defined ‘persistent symptoms’ as a child having either answered yes to the above question in the long covid survey, or reporting symptom episodes lasting 4 weeks or more through the weekly surveys. If the date of onset of persistent symptoms was missing from the long covid survey, the 20^th^ January 2021 was used as the onset date (5 weeks before the survey date). If information on persistent symptoms was derived from the weekly survey, the start date of the illness episode was used as the onset of persistent symptoms.

We coded persistent symptoms into groups used by the National Institute for Health and Care Excellence^8^: respiratory, cardiovascular, generalised (including fatigue and fever), neurological (including cognitive impairment/’brain fog’ and headache), gastrointestinal, psychological/psychiatric symptoms, ear, nose and throat (ENT) symptoms, dermatological or other symptoms.

Age was coded into three groups: <2, 2-11 and 12-17 years. Presence of a long-term condition was coded as a binary variable based on information regarding long-term conditions or medications from the baseline questionnaire. A small-area level indicator of socio-economic deprivation, the Index of Multiple Deprivation^9^ (IMD; coded into quintiles) and region of residence was mapped to the household postcode.

History of SARS-CoV-2 infection was defined where a child had i) reported a positive swab result, ii) had a positive swab as part of the VirusWatch survey, or iii) tested positive for SARS-CoV-2 IgG. If the estimated sample date of a positive swab result was prior to, or up to 10 days after the reported onset of persistent symptoms, we assumed children were infected before persistent symptom onset. The swab sample date was taken as the Monday in the week prior to the week of report of the positive sample for those individuals who had not had a sample collected as part of the VirusWatch study. For those reporting a positive swab test at baseline, only the month of the test was available; we estimated the sampling date to be the 15^th^ of the month. We assumed infection occurred prior to the start of persistent symptoms for IgG seropositive children.

We compared the distribution of age, sex, region of residence and IMD quintile in the study cohort with that of the resident population of children in England and Wales, derived from 2019 population estimates provided by the Office for National Statistics (ONS).^10 11^

We estimated the prevalence of persistent symptoms overall and in children with a history of SARS-CoV-2 infection according to age group, sex and presence of a long-term condition We fitted mixed effects logistic regression models for persistent symptom prevalence including these risk factors as the independent variables and household ID as the random intercept. We estimated the median duration of symptoms for children who had reported onset dates and that at least one symptom had ended. All analyses were carried out using Stata version 16 and RStudio version 3.4.3.

## Results

We included 4,678 children who met the inclusion criteria. Children aged 12-17 years were slightly over-represented in the VirusWatch child cohort compared to mid-year population estimates, as were children living in the Eastern regions of England. The child cohort was substantially less deprived than the population of children in England (Table 1).

**Table 1.** Characteristics of the VirusWatch Child Cohort and comparison with England & Wales population.

|  | **VirusWatch child cohort**<br>***n*=4,678** | **England & Wales** |
| --- | --- | --- |
| **Variable** | **Number (%)** |  |
| **Age group** |  |  |
| <2 | 329 (7.0) | 1,325,800 (10.5) |
| 2-11 years | 2522 (53.9) | 7,321,164 (57.9) |
| 12-17 years | 1827 (39.1) | 4,006,543 (31.7) |
| **Gender** |  |  |
| Male | 2091 (44.7) | 6,489,021 (51.3) |
| Female | 1898 (40.6) | 6,164,486 (48.7) |
| Missing | 689 (14.7) | - |
| **Region** |  |  |
| North East | 203 (4.3) | 532,057 (4.2) |
| North West | 479 (10.2) | 1,563,460 (12.4) |
| Yorkshire & Humber | 201 (4.3) | 1,169,941 (9.2) |
| East Midlands | 389 (8.3) | 1,002,649 (7.9) |
| West Midlands | 227 (4.9) | 1,299,803 (10.3) |
| East of England | 818 (17.5) | 1,346,457 (10.6) |
| London | 638 (13.6) | 2,032,427 (16.1) |
| South East | 817 (17.5) | 1,969,297 (15.6) |
| South West | 311 (6.7) | 1,107,477 (8.8) |
| Wales | 91 (1.9) | 629,939 (5.0) |
| Missing | 504 (10.8) | - |
| **IMD Quintile\*** |  |  |
| 1st quintile, most deprived | 371 (7.9) | 3,151,456 (23.7) |
| 2nd | 655 (14.0) | 2,753,699 (20.7) |
| 3rd | 832 (17.8) | 2,514,215 (18.9) |
| 4th | 1064 (22.7) | 2,402,419 (18.1) |
| 5th quintile, least deprived | 1252 (26.8) | 2,460,532 (18.5) |
| **Long term condition reported** |  |  |
| No | 4202 (89.8) | - |
| Yes | 476 (10.2) | - |
| **History of SARS-CoV-2 infection** |  |  |
| No | 4503 (96.3) | - |
| Yes | 175 (3.7) | - |
| **Persistent symptoms reported** |  |  |
| No | 4598 (98.3) | - |
| Yes | 80 (1.7) | - |

\*Population comparison group is children in England aged 0-19 years only

175 cohort children (3.7%) had evidence of past or present SARS-CoV-2 infection (Table 1). Of children with evidence of past infection, 110 had had a positive swab test (62.9%), 47 were positive on serology (26.9%) and 18 children had had a positive swab test and serology (10.3%). Seven children (of 175; 4%) had tested positive through the VirusWatch swabbing programme. Of the 476 children who reported at least one long-term condition, 385 (80.9%) reported having clinician-diagnosed asthma or using an inhaler (8.2% of the cohort).

The overall prevalence of persistent symptoms was 1.7% (80/4678 children; 95% CI 1.4%, 2.1%), and 4.6% (8/174 children; 95% CI 2.0%, 8.9%) in children who had a history of SARS-CoV-2 infection before persistent symptom onset.

Among children who reported persistent symptoms, the most common reported symptom types were general, ENT, and respiratory symptoms (figure 1). Among the 22 children who had reported at least one ‘general’ symptom, fatigue was the most common, reported by 18 children (22.5% of children reporting persistent symptoms).

**Figure 1.**
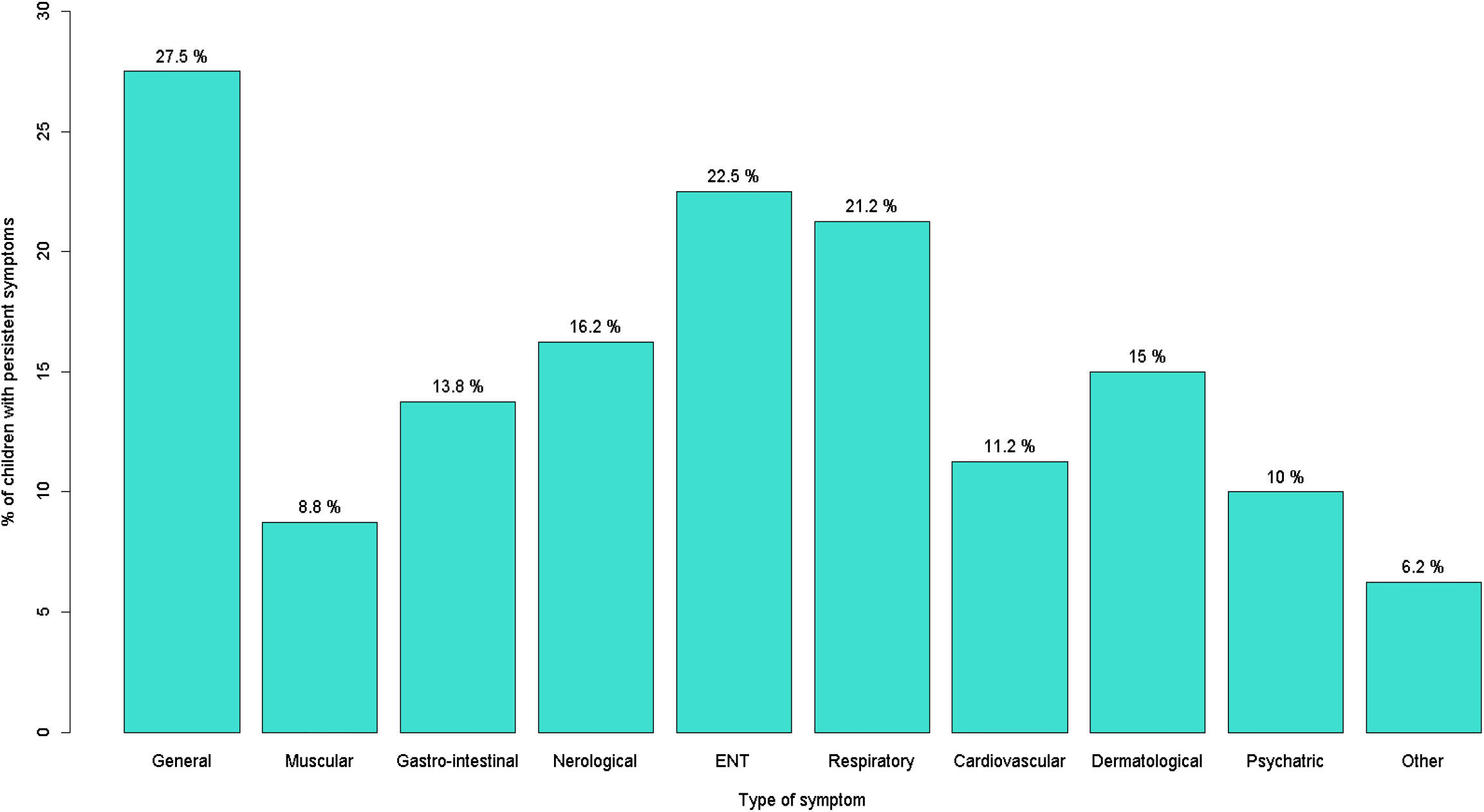
Distribution of types of symptoms among children reporting persistent symptoms (*n*=80)

The median duration of symptoms was 46 days (interquartile range 32-188) for the 18 children who reported start and end dates of symptoms.

Children who had evidence of SARS-CoV-2 infection were over twice as likely to report persistent symptoms compared to children who had not (Table 2). Being a teenager, girl or having long-term conditions significantly increased the odds of persistent symptoms.

**Table 2.**
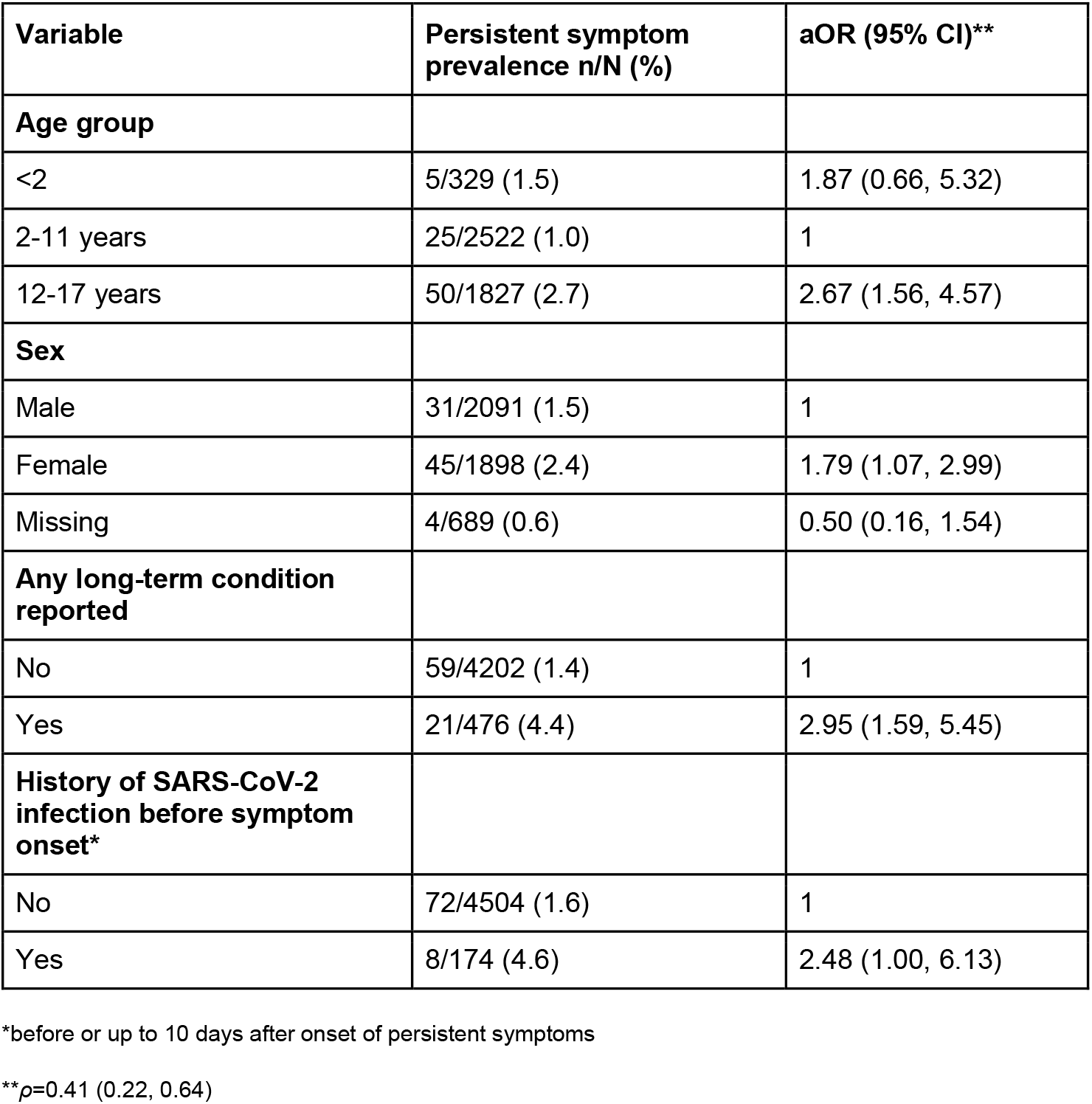
**Prevalence of persistent symptoms according to risk factors and adjusted odds ratios (aORs) from random effects logistic regression model**.

## Discussion

The prevalence of persistent symptoms lasting ≥4 weeks in children during the second and third UK wave of the COVID-19 pandemic was1.7% overall, and 4.6% among children with a history of SARS-CoV-2 infection. Apart from children with a history of SARS-CoV2 infection, girls, teenagers and children with long-term conditions were more likely to report persistent symptoms.

We used data from a large sample of children in England and Wales, which was representative of the general population of children in terms of age and sex, but less socio-economically deprived. VirusWatch collects swabs, blood samples and weekly reports of symptoms and test results. However, given that the prevalence of persistent symptoms was low, larger studies are required to assess risk factors for persistent symptoms in children related to SARS-CoV-2 infection in more detail. Since only one monthly survey to date have included questions on persistent symptoms, we were not able to assess time to symptom resolution for all children (as some had continuing symptoms), however evidence from other studies indicates that the majority of children recover after two months.^3 5^ Further, we were not able to compare the prevalence of persistent symptoms in children with SARS-CoV-2 infection to the proportion of children developing persistent symptoms after other respiratory infections.

Our estimate of the prevalence of persistent symptoms in children with a history of SARS-CoV-2 infection is similar to that reported via a UK symptom app,^3^ and in a study of primary school children with a history of SARS-CoV-2 infection,^12^ yet substantially lower than that reported in a UK-based survey.^4^ Notably, the prevalence in adults from the same survey was also substantially higher than reported elsewhere.^13^ Our estimated prevalence of persistent symptoms appears lower than among children seen in secondary care; nationally representative studies following children hospitalised with SARS-CoV-2 infection, including suitable control groups and using standardised definitions of persistent symptoms, are required in order to assess symptom persistence in children with more severe COVID-19 disease. The most common persistent symptom among children in Virus Watch was fatigue, as reported elsewhere.^3 5^

Whilst the prevalence of persistent symptoms in children was low in this study, most children reporting persistent symptoms did not have a known history of SARS-CoV-2 infection. Further research into persistent symptoms of SARS-CoV-2 infection in children to determine specific risk factors, and support for all children with persistent symptoms, irrespective of cause, are needed.

## Data Availability

We aim to share aggregate data from this project on our website and via a "Findings so far" section on our website - https://ucl-virus-watch.net/. We will also be sharing individual record level data on a research data sharing service such as the Office of National Statistics Secure Research Service. In sharing the data we will work within the principles set out in the UKRI Guidance on best practice in the management of research data. Access to use of the data whilst research is being conducted will be managed by the Chief Investigators (ACH and RWA) in accordance with the principles set out in the UKRI guidance on best practice in the management of research data. We will put analysis code on publicly available repositories to enable their reuse.

## Ethics

The Virus Watch study has been approved by the Hampstead NHS Health Research Authority Ethics Committee. Ethics approval number - 20/HRA/2320.

## Acknowledgements

We wish to thank all VirusWatch participants for their support for this study.

## Appendix 1

### Swab and antibody sampling and testing methods for Virus Watch

#### 1. Virus Watch PCR testing

Participants taking part in the laboratory testing cohort (please see the study protocol^1^ for further details regarding the Virus Watch laboratory testing cohort) were asked to submit nasopharyngeal swab specimens following certain symptom triggers suggestive of COVID-19 in themselves or a household member. Swab specimens were posted by the participants to the Francis Crick Institute, where they were tested for SARS-CoV-2 RNA via RT-PCR.

#### 2. Virus Watch serology testing

Children in Virus Watch were invited to provide blood samples for SARS-CoV-2 serology testing either at a clinic, where venous blood samples were taken via phlebotomy, or through a rapid diagnostic test using a finger prick capillary blood sample.

##### 2a. Clinic based blood sampling

A total of 10ml of whole blood were collected from children (≤15 years): by a member of the local research team with appropriate training and experience in phlebotomy. Children were offered Emla cream 20min before blood draw. Samples were sent to the Francis Crick Institute, where they were tested for IgG antibodies against the S1-subunit of the Spike protein of SARS-CoV-2.^2^

##### 2b. Finger prick blood sampling

Finger prick capillary blood samples were collected via self-sampling (aided by a parent/guardian for children), and participants used a rapid diagnostic test (Fortress Diagnostics, Antrim, UK) to self-test for SARS-CoV-2 IgG and IgM against the Spike protein.

## References

1. The Lancet. Facing up to long COVID. The Lancet 2020;396(10266):1861. doi: 10.1016/S0140-6736(20)32662-3

2. National Institute for Health Research. Living with COVID-19: Second Review, 2021. https://evidence.nihr.ac.uk/themedreview/living-with-covid19-second-review/; accessed 28/05/2021

3. Molteni E, Sudre CH, Canas LS, et al. Illness duration and symptom profile in a large cohort of symptomatic UK school-aged children tested for SARS-CoV-2. medRxiv 2021:2021.05.05.21256649. doi: 10.1101/2021.05.05.21256649

4. Office for National Statistics. All data relating to prevalence of ongoing symptoms following coronavirus (COVID-19) infection in the UK 2021 https://www.ons.gov.uk/peoplepopulationandcommunity/healthandsocialcare/conditionsanddiseases/datasets/alldatarelatingtoprevalenceofongoingsymptomsfollowingcoronaviruscovid19infectionintheuk accessed 10/05/2021.

5. Say D, Crawford N, McNab S, et al. Post-acute COVID-19 outcomes in children with mild and asymptomatic disease. The Lancet Child & Adolescent Health doi: 10.1016/S2352-4642(21)00124-3

6. Osmanov IM, Spiridonova E, Bobkova P, et al. Risk factors for long covid in previously hospitalised children using the ISARIC Global follow-up protocol: A prospective cohort study. medRxiv 2021:2021.04.26.21256110. doi: 10.1101/2021.04.26.21256110

7. Hayward A, Fragaszy E, Kovar J, et al. Risk factors, symptom reporting, healthcare-seeking behaviour and adherence to public health guidance: protocol for Virus Watch, a prospective community cohort study. medRxiv 2020:2020.12.15.20248254. doi: 10.1101/2020.12.15.20248254

8. National Institute for Health and Care Excellence. COVID-19 rapid guideline: managing the long-term effects of COVID-19, 2020. https://www.nice.org.uk/guidance/ng188/resources/covid19-rapid-guideline-managing-the-longterm-effects-of-covid19-pdf-66142028400325, accessed 28/05/2021

9. Ministry of Housing Communities and Local Government. The English Indices of Deprivation 2019 2019. https://www.gov.uk/government/statistics/english-indices-of-deprivation-2019 accessed 28/05/2021

10. Office for National Statistics. Populations by Index of Multiple Deprivation, England, 2001 to 2019 2020, https://www.ons.gov.uk/peoplepopulationandcommunity/populationandmigration/populationestimates/adhocs/12386populationbyindexofmultipledeprivationimdengland2001to2019 accessed 10/05/2021.

11. Office for National Statistics. Population projections by single year of age 2018, https://www.nomisweb.co.uk/datasets/ppsyoala accessed 10/05/2021.

12. Powell AA, Amin-Chowdhury Z, Mensah A, et al. Severe Acute Respiratory Syndrome Coronavirus 2 Infections in Primary School Age Children After Partial Reopening of Schools in England. Pediatr Infect Dis J 2021;40(6):e243–e45. doi: 10.1097/inf.0000000000003120 [published Online First: 2021/04/27]

13. Sudre CH, Murray B, Varsavsky T, et al. Attributes and predictors of Long-COVID: analysis of COVID cases and their symptoms collected by the Covid Symptoms Study App. medRxiv 2020:2020.10.19.20214494. doi: 10.1101/2020.10.19.20214494

## References

1. Hayward A, Fragaszy E, Kovar J, et al. Risk factors, symptom reporting, healthcare-seeking behaviour and adherence to public health guidance: protocol for Virus Watch, a prospective community cohort study. medRxiv 2020:2020.12.15.20248254. doi: 10.1101/2020.12.15.20248254

2. Ng KW, Faulkner N, Cornish GH, et al. Preexisting and de novo humoral immunity to SARS-CoV-2 in humans. Science 2020;370(6522):1339–43. doi: 10.1126/science.abe1107

